# After NICU Discharge: Preterm Infant Feeding and Growth Through the First Year

**DOI:** 10.1101/2022.10.03.22280654

**Authors:** Cristina R. Fernández

**Affiliations:** Columbia University Vagelos College of Physicians and Surgeons and NewYork-Presbyterian Hospital, New York, NY, USA

**Keywords:** very preterm infant, moderate preterm infant, post-hospital discharge feeding, infant growth, obesity, community pediatric provider

## Abstract

**Background:** Little is known about preterm infant feeding and growth in the outpatient community setting and there are no standardized post-hospital discharge feeding guidelines.

**Objective:** To describe the post-neonatal intensive care unit (NICU) discharge growth trajectories of very preterm (<32 weeks gestational age (GA)) and moderately preterm (32-34 6/7 weeks GA) infants managed by community pediatric providers and to determine the association between post-discharge feeding type and growth Z-scores and z-score changes through 12 months corrected age (CA).

**Methods:** Very preterm infants (n=104) and moderately preterm infants (n=109) born 2010-2014 and followed in hospital-affiliated community health clinics were enrolled in a single-center retrospective cohort study. Infant home feeding type and anthropometry were abstracted from outpatient medical chart review, and repeated measures analysis of variance calculated adjusted growth z-scores and z-score differences between 4 and 12 months CA. Linear regression models tested the relationship between type of home feeding at 4 months CA and anthropometry at 12 months CA.

**Results:** By 12 months CA, 5% of very preterm and 10% of moderately preterm infants were overweight. Moderately preterm infants on nutrient-enriched vs. standard term feeds had lower length z-scores at 12 months CA (−0.04 vs. 0.37, P=.03). Feeding type at 4 months CA predicted 12 month CA body mass index z-scores for very preterm infants (β=-0.66(−1.28, -0.04)).

**Conclusion:** Community pediatric providers may manage preterm infant post-NICU discharge feeding in the context of growth. Further research is needed to explore modifiable drivers of obesity risk in preterm infants to optimize infant development.

## BACKGROUND

Preterm birth, defined as birth less than 37 weeks gestational age (GA) affects 1 out of every 10 infants in the U.S [1]. Preterm birth has been associated with long-term health morbidity, such as neurocognitive impairment and obesity [2-4]. Obesity in children with history of preterm birth is thought to be related to rapid growth in the first years of life and has implications for chronic disease later in adulthood [5-8]. Understanding and interpreting preterm infant growth patterns and nutritional needs is crucial to support optimal early childhood development for this high-risk population. Yet less is known about how primary care pediatric providers are managing preterm infant feeding and growing in the months after neonatal intensive care unit (NICU) discharge.

Many preterm infants are considered growth-restricted by the time of hospital discharge. The majority of these preterm infants are discharged home with nutrient-enriched dietary feeding plans with milk enriched with additional calories and protein and to support enhanced growth and limit neurodevelopmental impairments [9-10]. These feeding plans are comprised of fortified expressed human breastmilk for breastfed infants and commercially available nutrient-enriched formula milk for formula-fed infants. Pediatric providers in the outpatient setting then assume the responsibility for managing and monitoring preterm infant diet and growth. However, there are currently no globally standardized feeding guidelines for preterm infants after hospital discharge, and the optimal duration to continue an infant’s enriched diet in the outpatient setting is not fully understood.

There is inconsistent evidence to support preterm infant increased weight gain and linear growth on nutrient-enriched feeds compared to standard term formula and/or breast milk after NICU discharge [11]. Inconsistency in study conclusions may be due to variability in randomized control trial sample sizes, populations, locations, duration of enrichment, duration of the trial, and feeding methods and frequency [10-13]. Conflicting conclusions and variability in methodology present a challenge to primary care pediatric providers seeking evidence-based recommendations on preterm infant post-hospital discharge nutrition and growth monitoring. Describing primary care pediatric provider management of preterm infant feeding and growth in the community setting will inform future research into early child development and the modifiable risk factors for overweight and obesity in preterm infants after NICU discharge.

This study described and compared post-NICU discharge feeding type and growth (weight, length, and body mass index (BMI) z-scores) from 4 months to 12 months corrected age (CA) and examined the relationship between feeding type at 4 months CA and anthropometry at 12 months CA in a cohort of low-income urban preterm infants. Study infants were all discharged from the NICU on nutrient-enriched diets and followed by board-certified primary care pediatric attending providers and pediatric trainees, including interns and residents. The study hypothesized that there would be significant differences in growth z-scores and difference in growth z-scores between infants continued on nutrient-enriched feeds vs. infants transitioned to standard term feeds by 4 months CA.

## METHODS

### Study Population

A single-center retrospective cohort study of a preterm infants ≤34 weeks GA who were discharged on nutrient-enriched feeds from a New York City urban teaching hospital from 2010-2014 and followed in the hospital-affiliated community-based primary care clinics was conducted. Of the 1,773 preterm infants ≤ 34 weeks GA discharged from the NICU from January 2010-December 2014, there were 279 infants (16%) discharged on nutrient-enriched feeds who followed in the hospital-affiliated primary care clinics (Figure 1). Of these 279 infants, only 213 infants were included in the analytic sample due to having complete anthropometry documented through 12 months CA. Infants were excluded from the study if they were discharged from the hospital NICU on human milk or standard full-term formula diets only; had systematic diseases that affected weight gain and required outpatient management by cardiology, gastroenterology, and/or pulmonology specialists; or were lost to primary care follow-up or had prior to reaching 12 months CA. The hospital-affiliated primary care clinics only care for children with public insurance for low-income families (such as Medicaid) or the New York State Children’s Health Insurance Program plan for low-income children. As moderate and late preterm infants account for the majority of preterm births in the U.S. [14], the infants in this study were categorized as either very preterm if GA was <32 weeks or as moderately preterm if GA was 32 to 34 weeks. Preterm infant categories were based on subgroups proposed by the U.S. National Institute of Child Health and Human Development [15]. Infant CA was defined as the menstrual age plus chronological age.

**Figure 1.**
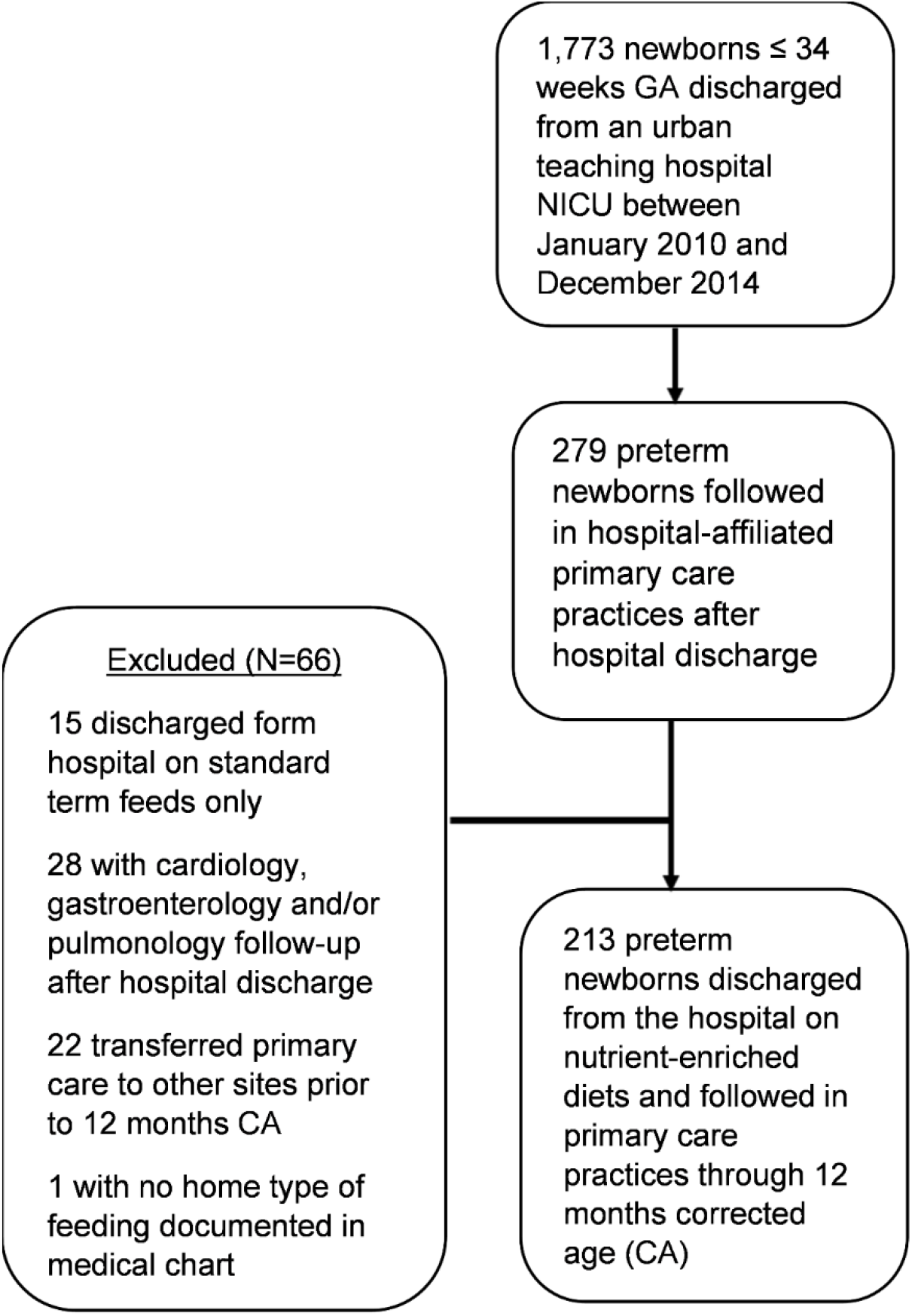
Study cohort selection flow diagram.

Infants who were lost to follow-up were not significantly different by gestational age, birth weight, sex, or ethnicity compared to infants who continued to follow at the primary care clinics through 12 months CA.

### Measures

The primary predictor was type of milk feeding at 4 months CA, as primary care providers are accustomed to timing of solid food introduction of 4-6 months for term infants [16]. Type of infant home feeding at 4 months CA was categorized as either nutrient-enriched feeds or exclusively standard term feeds. Nutrient-enriched feeds included post-hospital discharge formula with a minimum caloric density of 22 kcal/oz only or post-hospital discharge formula and human breastmilk. Standard term feeds included full-term infant formula with caloric density of 20 kcal/oz only, full-term formula and human breastmilk, or human breastmilk only. Information on formula manufacturers, volume and frequency of formula consumption, daily breastfeeding frequency, time of introduction of solid food, and daily volume and frequency of solids and juice intake were not available for analysis due to high variability in pediatrician documentation.

The primary outcomes were weight-for-age z-score, length-for-age z-score, and BMI-for-age z-score at 12 months and the difference in z-scores between 4- and 12-months CA by gestational age group and type of home feeding at 4 months CA. Weight (in kilograms (kg)) and length (to the nearest 0.5 centimeter (cm)) of infants were measured according to standard hospital and clinic procedures. BMI was calculated from the documented length and weight in the medical chart at the corrected age of interest. Z-scores for birth and discharge weight and length were calculated using the validated sex-specific Fenton Preterm Growth Charts [17]. Birth BMI z-scores and small for gestational age (SGA) status at birth were calculated using the validated Olsen 2015 BMI curves for preterm infants [18]. All growth z-scores at 4 months CA and 12 months CA were calculated using the World Health Organization (WHO) Growth Standards (WHO Anthro® 3.2.2, January 2011). Use of WHO Growth Standards are recommended for all children less than 24 months by the U.S. Centers for Disease Control [19]. BMI z-scores greater than +2.0 at 12 months CA were considered overweight [20].

All data were abstracted from the electronic medical record by the research team based on provider documentation during infant clinical visits. The data abstraction process was partly informed by a medical record abstraction framework proposed by Zozus et al. [21]. The research team created a hypothesis, reviewed the literature to operationalize measures, created a codebook to guide abstraction methods, and conducted team abstraction reviews to share feedback on the accuracy of abstraction and make adjustments as needed. The Institutional Review Board for Human Investigations at the Columbia University Irving Medical Center gave ethical approval for this study protocol with waiver of informed consent.

### Statistical Analyses

Comparison of descriptive characteristics between very preterm and moderately preterm infants were conducted using Chi-square and Fisher exact tests for categorical variables and Student’s t tests and Wilcoxon Rank Sum tests for continuous variables. Given the unbalanced group sizes of infants on nutrient-enriched vs. standard term feeds at 4-months CA, least square (adjusted) means for growth z-scores at birth, hospital discharge, 4 months CA, and 12-months CA were estimated for infants by gestational age group and type of feeding at 4 months CA, adjusted for birth z-score parameter. Differences in adjusted means for growth z-scores between 4 and 12 months CA in infants who were transitioned to standard term feeds vs. infants who were maintained on nutrient-enriched feeds by 4 months CA in the very preterm group and in the moderately preterm group were compared for very preterm and moderately preterm infant groups. Difference in mean growth from 4 to 12 months CA between infants on nutrient-enriched feeds vs. standard term feeds at 4 months were quantified for the very preterm and moderately preterm infant groups using repeated measures analysis of variance (ANOVA), controlling for birth parameter z-score. Linear regression models estimated the relationship between type of feeding at 4 months CA and anthropometry at 12 months CA in the entire sample, adjusting for birth z-score parameter. SAS® software (Version 9.4, SAS Institute, Inc.) was used for all statistical analyses. Statistical tests were 2-sided and statistical significance was carried out at the P < .05 level.

## RESULTS

Of the 213 infants in the analytic sample, n=104 were very preterm and n=109 were moderately preterm. Characteristics of infants are available in Table 1. Very preterm infants had significantly lower birth weight (1127 ± 370 g vs. 1994 ± 432 g, respectively), longer hospital stay (61± 28 days vs. 17 ± 11 days, respectively), and an older corrected age at hospital discharge (37.1 ± 2.7 vs. 35.6 ± 1.3, respectively). There were no significant differences between the preterm infant groups by maternal age or mode of delivery, sex, Latino ethnicity, small for gestational age at birth status, and mean number of emergency room visits within the first year of life. About 25% of very preterm infants compared to 50% of moderately preterm infants were also breastfeeding at NICU discharge (P=.0002), and only 4% of very preterm infants and 6% of moderately preterm infants were still breastfeeding by 4-months CA (P=.80). Eighty-seven percent of very preterm infants and 72% of moderately preterm infants were continued on nutrient-enriched feeds at 4 months CA. By comparison, 13% of very preterm infants and 28% of moderately preterm infants who were transitioned to standard term feeds by their pediatrician by 4 months CA (P=.009). None of the very preterm infants and 21% of the moderately preterm infants were transitioned from nutrient-enriched feeds to standard term feeds between 4 months and 6 months CA. Five percent (n=5) of very preterm infants and 8% (n=11) of moderately preterm infants had BMI z-scores greater than +2.0 considered overweight at 12 months CA.

**Table 1.** Characteristics of Very Preterm (<32 weeks GA) and Moderately Preterm (32-34 weeks GA) Infants (N=213)

| Characteristic | Very preterm infants<br>(n= 104) | Moderate preterm<br>infants<br>(n=109) | *p* |
| --- | --- | --- | --- |
| Mean maternal age at delivery (years) | 28.9 ± 6.0 | 24.4 ± 6.2 | .51 |
| Parity | 1.8 ± 0.9 | 2.3 ± 1.2 | .20 |
| Cesarean delivery | 71 (68) | 71 (65) | .07 |
| Sex, *n* (%) |  |  | .33 |
| Male | 47 (46) | 57 (52) |  |
| Female | 56 (54) | 52 (47) |  |
| Mean gestational age (GA) | 28 3/7 | 33 2/7 | < .00001 |
| Latino ethnicity, *n* (%) | 89 (86) | 87 (80) | .16 |
| Birth weight (g) | 1126.7 ± 370.0 | 1994.2 ± 431.9 | <.0001 |
| Birth weight z-score | 0.004 ± 1.0 | -0.09 ± 0.9 | .44 |
| Birth length (cm) | 37.12 ± 2.8 | 43.9 ± 2.8 | <.0001 |
| Birth length z-score | 0.21 ± 1.1 | 0.2 ± 0.9 | .93 |
| Birth BMI (kg/m<sup>2</sup>) | 8.0 ± 1.3 | 10.2 ± 1.5 | <.0001 |
| Birth BMI z-score | -0.15 ± 1.2 | -0.03 ± 1.2 | .47 |
| Small for gestational age, *n* (%) | 9 (9) | 9 (9) | .90 |
| Total hospital stay (days) | 61.2 ± 28.4 | 16.6 ± 10.6 | <.0001 |
| Corrected age at hospital discharge<br>(weeks) | 37.1 ± 2.7 | 35.6 ± 1.3 | <.0001 |
| Hospital discharge weight z-score | -0.83 ± 1.3 | -0.61 ± 0.8 | .29 |
| Hospital discharge length z-score | -0.74 ± 1.2 | -0.12 ± 0.9 | .0001 |
| Hospital discharge BMI z-score | -0.14 ± 1.6 | -0.42 ± 0.9 | .03 |
| Breastfeeding at discharge, *n* (%) | 26 (25) | 55 (50) | .0002 |
| Breastfeeding at 4 months CA, *n* (%) | 4 (4) | 6 (6) | .80 |
| Type of home feeding at 4 months CA |  |  | .009 |
| Nutrient-enriched | 88 (87) | 79 (72) |  |
| Standard term | 13 (13) | 30 (28) |  |
| Mean emergency room visits by 12 months CA | 2.5 ± 2.2 | 2.0 ± 2.1 | .08 |

The adjusted mean growth z-scores at birth, hospital discharge, 4 months CA, and 12 months CA for infant GA groups stratified by type of home feeding at 4 months CA are visualized in Figure 2 (length z-scores), Figure 3 (weight z-scores), and Figure 4 (BMI z-scores). Moderately preterm infants on nutrient-enriched feeds had significantly lower adjusted mean weight [-0.26 (0.11) vs. 0.34 (0.18), P=.005] and length z-scores at 4 months CA compared to infants on standard term feeds [-0.35 (0.13) vs. 0.10 (0.21), P=.03 respectively]. Moderately preterm infants on nutrient-enriched feeds also had significantly lower adjusted mean length z-scores at 12 months CA [-0.04 (0.13) vs. 0.37 (0.21), respectively, P=.03] compared to moderately preterm infants on standard term feeds. There were no significant differences observed in the adjusted mean growth z-scores at 4 and 12 months CA in very preterm infants receiving nutrient-enriched vs. standard term feeds at 4 months CA.

**Figure 2.**
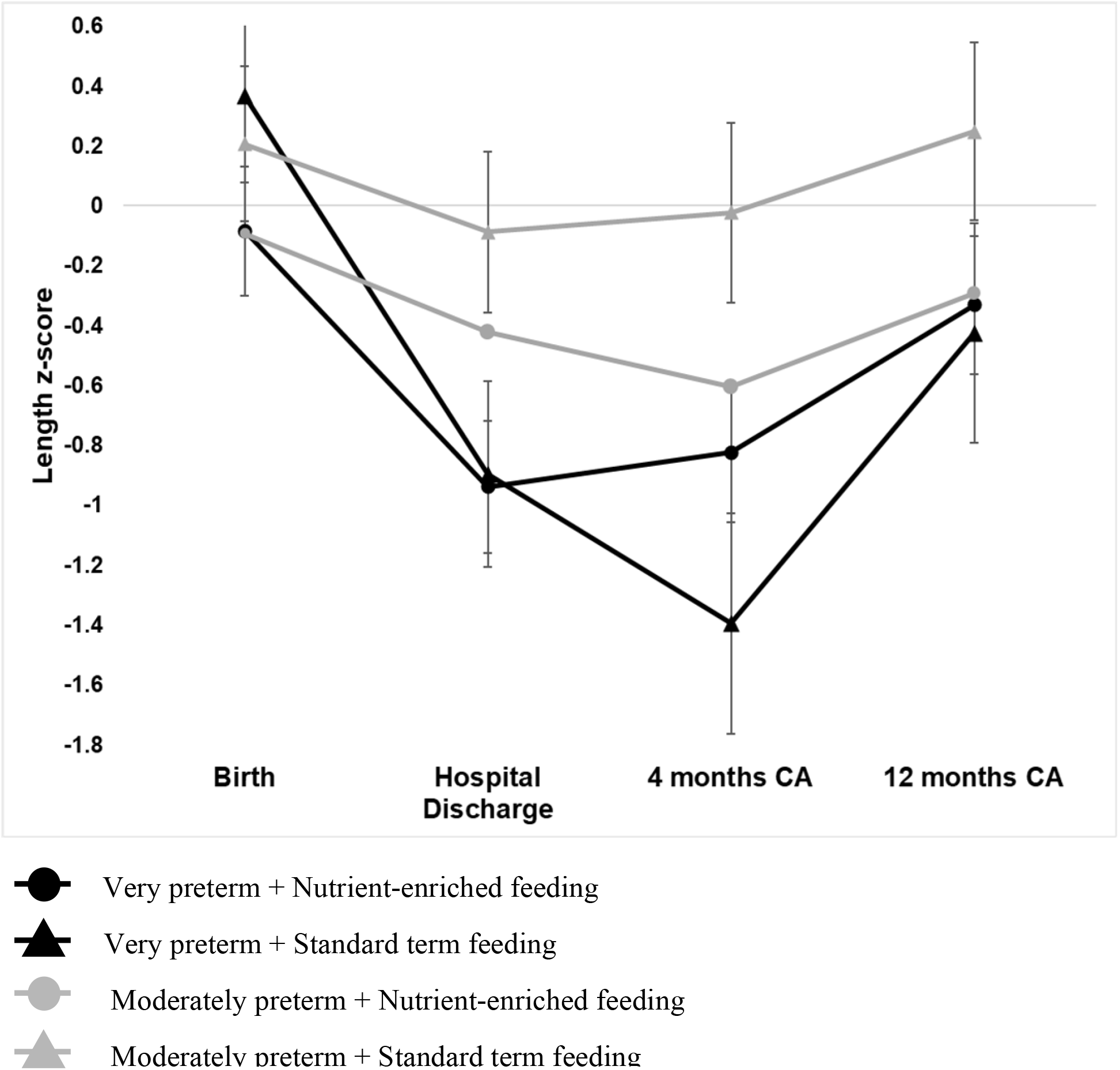
Adjusted mean length z-scores of infants through 12 months corrected age (CA) by type of home milk feeding at 4 months CA and by gestational age. Length z-scores are displayed as adjusted mean z-scores ± standard error of means from repeated measures analysis of variance with an unstructured covariance matrix was performed, adjusted for birth length z-score. Moderately preterm infants on enriched feeds had persistently lower adjusted mean length z-scores at 4 and 12 months CA compared to moderately preterm infants on standard term feeds, although the difference was not statistically significant by 12 months CA.

**Figure 3.**
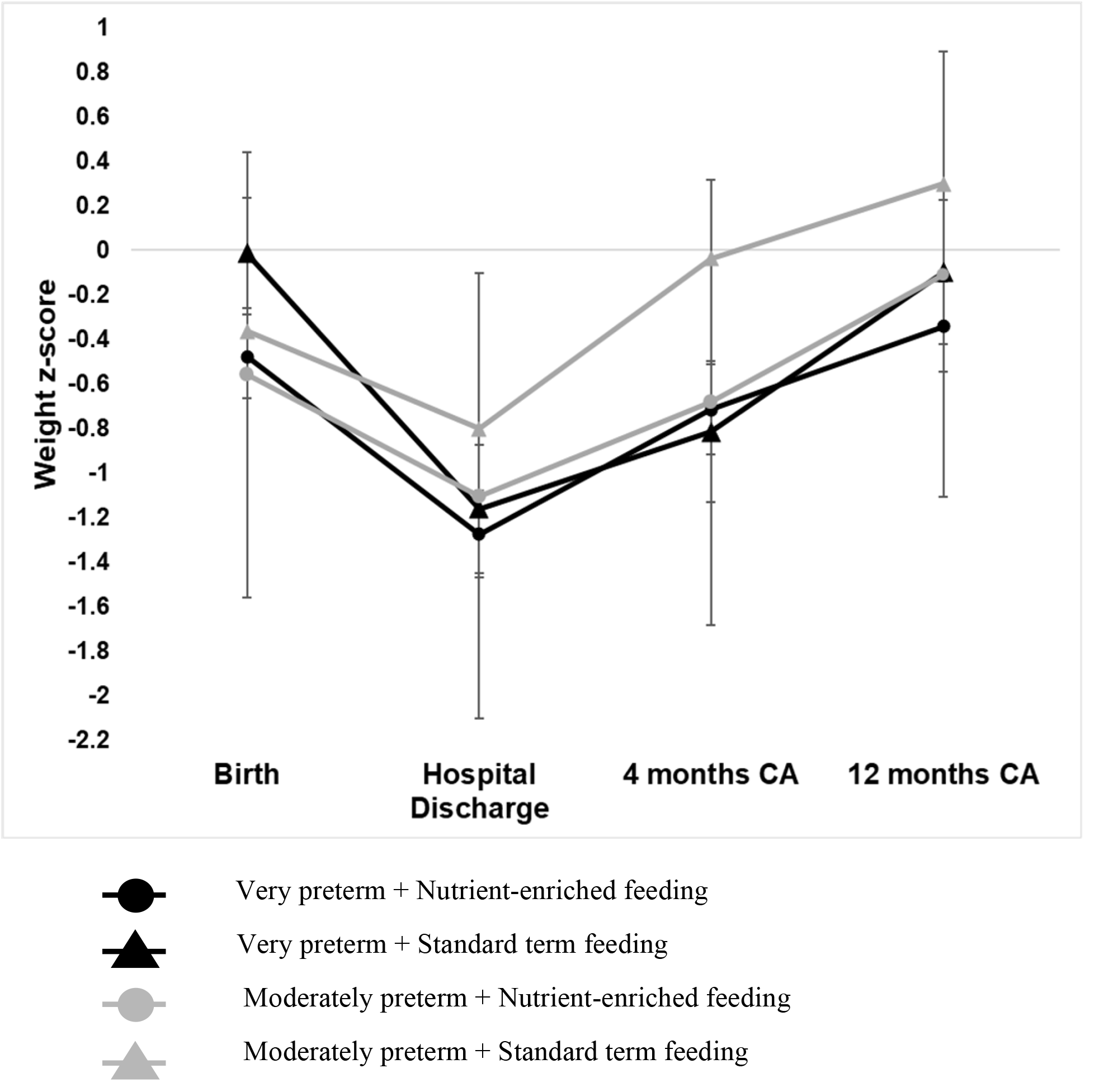
Adjusted mean weight z-scores through 12 months corrected age by feeding type at 4 months corrected age. Adjusted mean weight z-scores of infants through 12 months corrected age (CA) by type of home milk feeding at 4 months CA and by gestational age. Weight z-scores are displayed as adjusted mean z-scores ± standard error of means from repeated measures analysis of variance with an unstructured covariance matrix was performed, adjusted for birth weight z-score. Moderately preterm infants on nutrient-enriched feeds had significantly lower adjusted mean weight z-scores at 4 months CA compared to moderately preterm infants on standard term feeds.

**Figure 4.**
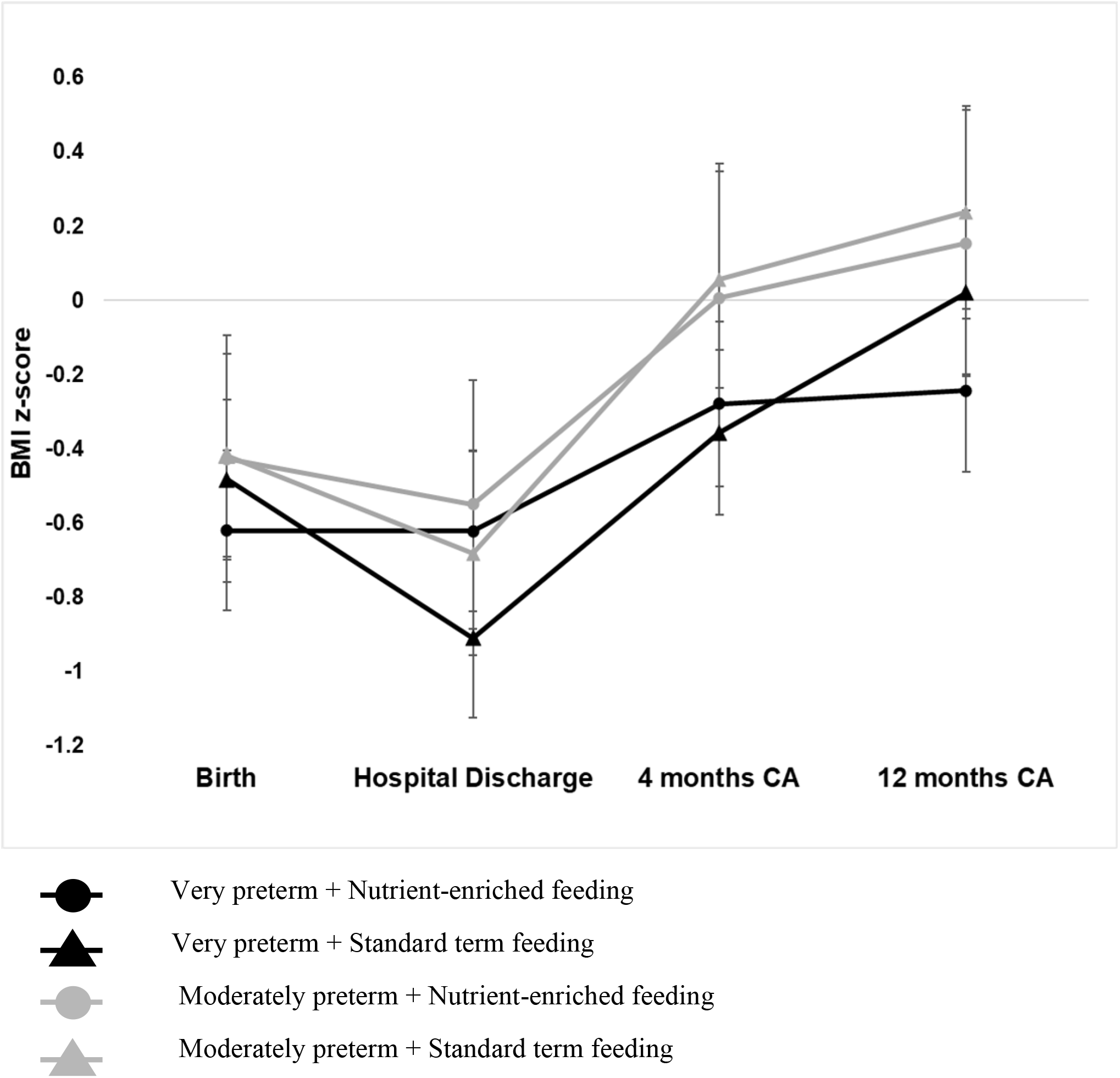
Adjusted mean body mass index z-scores through 12 months corrected age by feeding type at 4 months corrected age. Adjusted mean body mass index (BMI) z-scores of infants through 12 months corrected age (CA) by type of home milk feeding at 4 months CA and by gestational age. BMI z-scores are displayed as adjusted mean z-scores ± standard error of means from repeated measures analysis of variance with an unstructured covariance matrix was performed, adjusted for birth BMI z-score. There was no significant difference in adjusted mean BMI z-score between very preterm infants on nutrient-enriched feeds vs. standard term feeds. There was no significant difference in adjusted mean BMI z-score between moderately preterm infants on nutrient-enriched feeds vs. standard term feeds.

Very preterm infants transitioned to standard term feeds by 4 months CA had significantly higher adjusted weight z-score (Δweight= -0.37, P<.0001) and length z-score (Δlength= -0.97, P=.003) at 12 months CA compared to 4 months CA (Table 2). Similarly, very preterm infants maintained on nutrient-enriched feeds through 4 months CA had significantly higher adjusted weight (Δweight= -0.37, P<.0001) and length (Δlength= -0.49, P<.0001) z-scores at 12 months CA compared to 4 months CA. Moderately preterm infants transitioned to standard term feeds by 4 months CA had a significantly higher adjusted weight z-score at 12 months CA compared to 4 months CA (Δweight= -0.33, P=.02) (Table 3). Moderately preterm infants maintained on nutrient-enriched feeds through 4 months CA had significantly higher adjusted weight (Δweight= -0.58, P<.0001) and BMI z-scores at 12 months CA (ΔBMI= -0.34, P=.03). There was no significant difference in the magnitude of the change in z-score for weight, length, or BMI for either very preterm infants or moderately preterm infants when comparing infants transitioned to standard term feeds vs. maintained on nutrient-enriched feeds through 4 months CA.

**Table 2.**
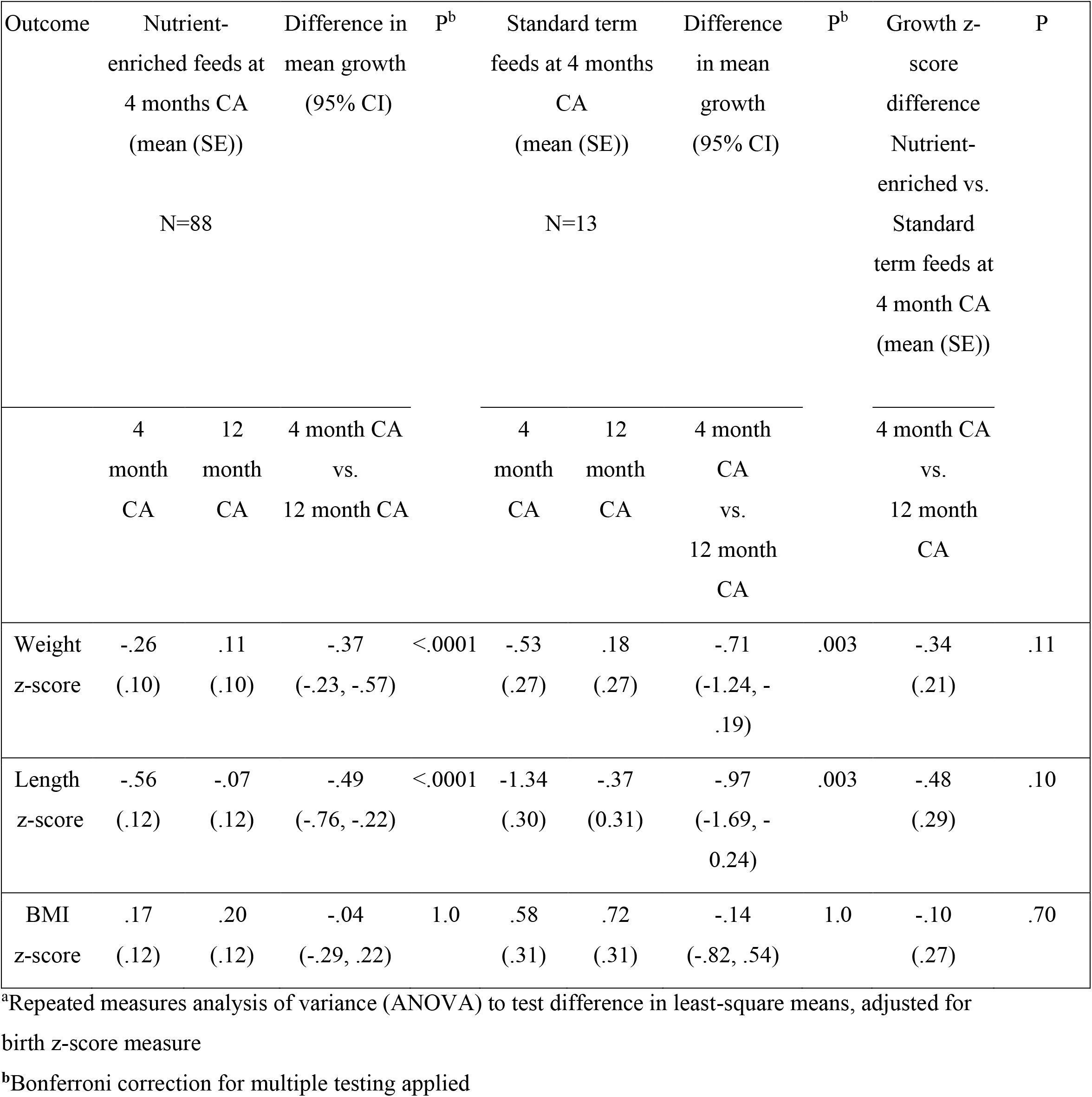
Adjusted^a^ Growth Z-scores and Growth Z-score Difference at 4 Months vs. 12 Months Corrected Age (CA) for Very Preterm Infants, by Feeding Type

**Table 3.** Adjusted^a^ Growth Z-scores and Growth Z-score Difference at 4 Months vs. 12 Months Corrected Age (CA) for Moderately Preterm Infants, by Feeding Type

| Outcome | Nutrient-enriched feeds at 4 months CA<br>(mean (SE))<br>N=79 |  | Difference in mean growth (95% CI) | p<sup>b</sup> | Standard term feeds at 4 months CA<br>(mean (SE))<br>N=29 |  | Difference in mean growth (95% CI) | p<sup>b</sup> | Growth z-score difference Nutrient-enriched vs. Standard term feeds at 4 months CA<br>(mean (SE)) | p |
| --- | --- | --- | --- | --- | --- | --- | --- | --- | --- | --- |
|  | 4 months CA | 12 months CA | 4 months CA vs. 12 months CA |  | 4 months CA | 12 months CA | 4 months CA vs. 12 months CA |  | 4 months CA vs. 12 months CA |  |
| Weight z-score | -.26<br>(.11) | .32<br>(.11) | -.58<br>(-.81, -.34) | <.0001 | .34<br>(.18) | .68<br>(.18) | -.33<br>(-.60, -.06) | .02 | .24<br>(0.17) | .16 |
| Length z-score | -.35<br>(.13) | -.04<br>(.13) | -.30<br>(-.61, .007) | .06 | .10<br>(.21) | .37<br>(.21) | -.27<br>(-.78, .23) | .88 | .03<br>(.22) | .90 |
| BMI z-score | .08<br>(.14) | .42<br>(.14) | -.34<br>(-.65, -.03) | .03 | .51<br>(.22) | .68<br>(.22) | -.17<br>(-.68, .33) | 1.0 | .16<br>(.22) | .46 |

<sup>a</sup>Repeated measures analysis of variance (ANOVA) to test difference in least-square means, adjusted for birth z-score measure
<sup>b</sup>Bonferroni correction for multiple testing applied

Results of adjusted linear regression models estimating the association between type of home milk feeding and 12-month CA weight, length, and BMI z-scores are reported in Table 4 for very preterm infants and Table 5 for moderately preterm infants. Type of home feeding at 4 months CA significantly predicted 12-month CA BMI z-scores in very preterm infants (β= **-0.66 (−1.28, - 0.04)**, P=.03), adjusting for birth z-score parameter.

**Table 4.** Association of Type of Home Feeding at 4 Months CA with Anthropometry at 12 Months Corrected Age in Very Preterm Infants, Adjusted for Birth Z-score Parameter

|  | **Weight Z-score** | **Length Z-score** | **Body Mass Index Z-score** |
| --- | --- | --- | --- |
| **Variable** |  |  |  |
| **Home feeding practices at 4 months corrected age (reference: standard term)** | **-.19 (-.26, .84)<sup>a</sup>** | **.27 (-.40, .94)** | **-.66 (-1.28, -.04)\*** |
| **Birth z-score parameter** | **.50 (.30, .69)\*\*** | **.50 (.31, .70)\*\*** | **-.08 (-.26, .09)\*\*** |

<sup>a</sup>Values reported as adjusted standardized $\beta$ coefficient (95% confidence interval)
\* $p < .05$ ; \*\* $p < .01$

**Table 5.** Association of Type of Home Feeding at 4 Months CA with Anthropometry at 12 Months Corrected Age in Moderately Preterm Infants, Adjusted for Birth Z-score Parameter

|  | Weight Z-score | Length Z-score | Body Mass Index Z-score |
| --- | --- | --- | --- |
| Variable |  |  |  |
| Home feeding practices at 4 months corrected age (reference: standard term) | -.37 (-.81, .06)<sup>a</sup> | -.49 (-1.0, .01) | -.27 (-.83, .27) |
| Birth z-score parameter | .42 (.20, .64)** | .38 (.13, .63)** | .26 (.06, .46)* |

<sup>a</sup>Values reported as adjusted standardized $\beta$ coefficient (95% confidence interval)
\*p<.05; \*\*p<.01

## DISCUSSION

In this sample of urban low-income very preterm and moderately preterm infants in hospital-affiliated primary care clinics, the majority of infants were on nutrient-enriched feeds through at least 4 months CA. However, both very preterm infants and moderately preterm infants demonstrated significant weight z-score increases from 4 months CA to 12 months CA, irrespective of type of feeding at 4 months CA. Very preterm infants had no significant differences in adjusted weight, length, or BMI z-scores at 4 months CA or 12 months CA between nutrient-enriched and standard term feed groups. This suggests that primary care pediatricians may have considered additional factors beyond gestational age and infant growth measurement when managing preterm infant home feeding.

By contrast, moderately preterm infants maintained on nutrient-enriched feeds through at least 4 months CA had a persistently significantly lower adjusted length z-score from 4 months CA to 12 months CA compared to infants transitioned to standard term feeds at 4 months CA. Whether this significant difference in length z-scores is clinically meaningful remains a question for further research given the continued global conversation on ideal curves to monitor linear growth of preterm infants after hospital discharge [17,22,23]. While nutrient-enriched feeds have an increased amount of protein thought to contribute to greater linear growth [24], non-nutritional factors may also influence linear growth of preterm infants. This is especially pertinent given that moderately preterm infants on nutrient-enriched feeds at 4 months CA had no significant difference in adjusted weight z-score at 12 months compared to infants transitioned to standard term feeds at 4 months CA.

Type of feeding at 4 months CA significantly predicted 12 months CA mean BMI z-scores in very preterm infants when adjusting for birth z-score parameter. Further research quantifying preterm infant home feeding frequency and volume through at least 4 months CA will help provide pediatric primary care providers with a timeframe to re-evaluate infant home milk feeding and to consider ramifications on future growth.

There are unique challenges with the care of preterm infants, and the absence of standardized home feeding guidelines can increase difficulties faced by primary care pediatric providers in monitoring and managing preterm infant feeding and growth. A range in the awareness of breastfeeding guidelines for preterm infants was seen in a study of outpatient pediatricians in New York on challenges with preterm infant care after NICU discharge [25]. There was a decrease in breastfeeding rates after hospital discharge amongst mothers of both very preterm and moderately preterm infants. The results are consistent with those of a multiregional European study that found very preterm infants had a significantly lower probability of breastfeeding continuation if they received formula at discharge and their mothers were younger age and less educated [26]. Additionally, a study on breastfeeding cessation found that U.S. younger mothers and those with limited socioeconomic resources were more likely to stop breastfeeding within the first month after delivery [27]. Given that breastmilk is highly suggestive to reduce weight gain velocity and BMI in full-term infants [28], it is possible that continued breastfeeding in the study population may have decreased the weight z-scores seen at 12 months CA.

This study suggests an importance of supporting primary care pediatric providers’ efforts to educate families on the benefits of breastmilk for preterm infant growth after hospital discharge and to develop practice-based lactation support services targeting families of preterm infants [29], Forty-eight percent of community pediatricians caring for NICU graduates reported a lack of clinical experience with high-risk infants as an impedance to providing quality care to NICU graduates when surveyed in the Northeast U.S [30]. Community pediatric providers can support families who may have difficulty understanding the preterm infants’ discharge feeding plans, especially when mixing of formula powder and milk or water on hospital discharge instructions differs from the instructions on the formula container or human milk fortifier bag [31].

Pediatric primary care providers have many preterm infant growth charts, including the Fenton 2013 and the INTERGROWTH-21^st^ Preterm Postnatal Growth Standard 2015, that can be used to monitor growth and guide nutritional management up to term or post-term through about 3-6 months CA [18,22,32]. The growth monitoring tool selected by the pediatric primary care provider can depend on the provider’s preference and local norms, with ramifications for provider assessment on normal or abnormal preterm infant growth [23]. Use of birthweight-for-gestational age charts, constructed from cross-sectional birth size, to monitor growth over time may promote rapid growth to approximate intra-uterine fetal growth, with implications for future obesity risk [33-34]. Goal weight gain of g/kg/day or reaching the 5^th^-10^th^ percentile on the term infant growth chart to monitor preterm infant growth in the primary care setting may be limited by variability of periods of accelerated growth among different infants. Variability in birth weight, morbidity, gender and feeding can contribute to differing interpretations of rapid weight gain and catch-up growth. Lack of consensus on best methods to assess preterm infant growth in the primary care setting contributes to challenges comparing growth between infants when different methods are utilized.

Model instability from the small sample size of infants with overweight precluded an analysis of the association of post-discharge feeding type with overweight. Yet the presence of preterm infants with overweight raises implications for early obesity risk in a vulnerable population. Primary care pediatric providers should give consideration to risk for overweight and obesity as part of infant growth and milk feeding type management strategies [35].

There was no significant difference in the magnitude of the change in z-score for weight, length, or BMI between 4 months CA and 12 months CA for either very preterm infants or moderately preterm infants. This suggests the need for additional research with a larger sample of very preterm and moderately preterm infants to improve the power to detect a significant difference in 12 month adjusted mean growth z-scores. There are also likely additional factors, unmeasured in this study, that have previously been associated with preterm infant growth after hospital discharge, such as maternal factors (pre-pregnancy BMI, education, and smoking), household attitudes and beliefs around infant feeding, milk feeding volume and frequency, and preterm activity and sleep [24,36,37]. Mothers of preterm infants have reported experiencing anxiety and stress around identification of infant hunger and satiety cues, variability in infant feeding effectiveness, management of feeding volumes, and knowing how and when to advance feeds at home [38-39].

This study had several limitations. This was a single center, observational study that utilized a retrospective cohort. A causal relationship between type of home feeding at 4 months CA and 4 and 12 months CA growth measures could not be determined. While collecting data in one health care system was accessible and cost-effective, the results may not be generalizable across primary care sites and clinics. Type of home feeding and infant anthropometry were obtained from medical chart review rather than objectively collected or verified. Nutritional data could have been subject to recall bias or social desirability bias by caregivers wishing to appear to be following the primary care pediatricians’ nutrition management instructions. Reliance on secondary data may also have increased susceptibility to recall bias by pediatricians to accurately recording the infant home feeding at the follow-up visits.

Inconsistencies in primary care provider documentation of volume and frequency of home milk feeds and introduction and frequency of intake of complementary foods and beverages other than milk before 12 months CA precluded the use of this information in the analyses. These home feeding factors have also been shown to influence infant growth and may have contributed to the anthropometry findings at 12 months CA. Additionally, early growth in formula-fed preterm infants may be associated with specific composition of nutrient-enriched formula with regards to ratios of energy, protein, and mineral content [40]. Brand name of formula used by the infants’ families and formula composition information were not consistently documented and could not be used in analyses. Study limitations also included inconsistent information on maternal characteristics that have been associated with infant weight and growth as several infants were transferred to the urban teaching hospital NICU for management after their births at outside institutions. There may have been other unmeasured socio-environmental factors that had residual confounding effects on both infant feeding and growth, such as food and housing hardship experienced by the families and neighborhood activity spaces. In spite of these limitations, the study’s strengths included a sample of infants of largely Latino ethnicity from a low-income urban background, a population largely underrepresented in research. Additionally, a follow-up period for growth measures data extending to 12 months CA allowed us to capture the important time period between post-hospital discharge and late infancy.

This study contributes to the literature by characterizing the primary care pediatrician-managed home milk feeding and growth of urban, low-income very preterm and moderately preterm infants in the community in the months after hospital discharge. Future research aims to collect longitudinal quantitative and qualitative data on preterm infant home feeding frequencies and volumes, introduction of complementary foods, participation in the government nutrition assistance programs, and caregiver beliefs around preterm infant feeding and growing. Describing and comparing early infant and childhood growth patterns of preterm infants with different types of home milk feeding patterns has implications for primary care pediatric provider management of preterm infants after hospital discharge.

## CONCLUSIONS

A lack of consensus on post-hospital discharge feeding guidance can present a challenge to the primary care pediatric provider caring for preterm infants in the community setting. The study demonstrated that moderately preterm infants on enriched feeds had persistently lower length z-score at 4 and 12 months CA compared to moderately preterm infants on standard term feeds. Type of home feeding at 4 months CA predicted BMI z-score at 12 months CA for very preterm infants. Future work is needed to longitudinally investigate home milk feeding volume and frequency and to determine household and clinical factors that may contribute to primary care pediatric provider care of preterm infants to optimize their early development.

## Data Availability

The dataset used during the current study is available from the corresponding author upon reasonable request.

## ACKNOWLEDGEMENTS

I would like to thank Drs. Robert Whittington and Melissa Stockwell for their valuable comments on previous drafts of this manuscript.

